# Susceptibility to SARS-CoV-2 infection amongst children and adolescents compared with adults: a systematic review and meta-analysis

**DOI:** 10.1101/2020.05.20.20108126

**Authors:** Russell M. Viner, Oliver T. Mytton, Chris Bonell, G.J. Melendez-Torres, Joseph Ward, Lee Hudson, Claire Waddington, James Thomas, Simon Russell, Fiona van der Klis, Archana Koirala, Shamez Ladhani, Jasmina Panovska-Griffiths, Nicholas G. Davies, Robert Booy, Rosalind M. Eggo

**Author notes:** Corresponding author Prof. Russell Viner, UCL Great Ormond St. Institute of Child Health, 30 Guilford St. London WC1N 1EH, UK.

## Abstract

**Importance:** The degree to which children and young people are infected by and transmit the SARS-CoV-2 virus is unclear. The role of children and young people in transmission of SARS-CoV-2 is dependent on susceptibility, symptoms, viral load, social contact patterns and behaviour.

**Objective:** We undertook a rapid systematic review to address the question “What is the susceptibility to and transmission of SARS-CoV-2 by children and adolescents compared with adults?”

**Data sources:** We searched PubMed and medRxiv up to 28 July 2020 and identified 13,926 studies, with additional studies identified through handsearching of cited references and professional contacts.

**Study Selection:** We included studies which provided data on the prevalence of SARS-CoV-2 in children and young people (<20 years) compared with adults derived from contact-tracing or population-screening. We excluded single household studies.

**Data extraction and Synthesis:** We followed PRISMA guidelines for abstracting data, independently by 2 reviewers. Quality was assessed using a critical appraisal checklist for prevalence studies. Random effects meta-analysis was undertaken.

**Main Outcomes:** Secondary infection rate (contact-tracing studies) or prevalence or seroprevalence (population-screening studies) amongst children and young people compared with adults.

**Results:** 32 studies met inclusion criteria; 18 contact-tracing and 14 population-screening. The pooled odds ratio of being an infected contact in children compared with adults was 0.56 (0.37, 0.85) with substantial heterogeneity (95%). Three school contact tracing studies found minimal transmission by child or teacher index cases. Findings from population-screening studies were heterogenous and were not suitable for meta-analysis. The majority of studies were consistent with lower seroprevalence in children compared with adults, although seroprevalence in adolescents appeared similar to adults.

**Conclusions:** There is preliminary evidence that children and young people have lower susceptibility to SARS-CoV-2, with a 43% lower odds of being an infected contact. There is weak evidence that children and young people play a lesser role in transmission of SARS-CoV-2 at a population level. Our study provides no information on the infectivity of children.

**Key points:** *Question:* What is the evidence on the susceptibility and transmission of children and young people to SARS-CoV-2 in comparison with adults?

*Findings:* In this systematic review and meta-analysis, children and young people under 18-20 years had an 435 lower odds of secondary infection of with SARS-CoV-2 compared to adults 20 years plus, a significant difference. This finding was most marked in children under 12-14 years. Data were insufficient to conclude whether transmission of SARS-CoV-2 by children is lower than by adults.

*Meaning:* We found preliminary evidence that children have a lower susceptibility for SARS-CoV-2 infection compared with adults, although data for adolescents is less clear. The role that children and young people play in transmission of this pandemic remains unclear.

## Background

The degree to which children and young people under 20 years are infected by and transmit the SARS-CoV-2 virus is an unanswered question.^1-3^ These data are vital to inform national plans for relaxing social distancing measures including reopening schools.

Children and young people account for 1-3% of reported cases across countries^4-8^ and an even smaller proportion of severe cases and deaths.^5,9^ Children appear more likely to have asymptomatic infection than adults and analyses based upon symptom-based series underestimate infections in children.

The role that children and young people play in transmission of SARS-CoV-2 by is dependent upon their risk of exposure, their probability of being infected upon exposure (susceptibility), the extent to which they develop symptoms upon infection, the extent to which they develop a viral load sufficiently high to transmit and their propensity for making potentially infectious contact with others, dependent upon numbers of social contacts across age-groups and behaviour during those contacts.

Different study types may provide useful information on susceptibility and transmission in children compared with adults, yet each is open to bias. Contact-tracing studies with systematic follow-up of all contacts to estimate secondary attack rates (SAR) in children and adults can provide strong evidence on differential susceptibility. Findings from some contact tracing studies suggest that children have lower SARS-CoV-2 SAR than adults,^10^ although others have found no difference by age.^11^ One study from South Korea has suggested adolescents but not children may have higher SAR,^12^ although a separate analysis of child cases from the same population identified minimal transmission from these cases.^13^

Population-screening studies may identify infection through viral RNA detection or antibodies indicating prior infection. However the prevalence of SARS-CoV-2 in children in a population is not a direct indicator of susceptibility or transmission as the expected prevalence depends on exposure, susceptibility, proportions of children in the population, mixing rates among children and between adults and children and timing of social distancing interventions that disrupt mixing.

A number of authors have concluded that children and young people may be less susceptible to SARS-CoV-2,^2,14^ although there are multiple sources of bias in each study type which can complicate straightforward analysis. In contact-tracing studies, testing of only symptomatic contacts will introduce significant bias, as will seroprevalence studies drawn from clinical contact studies (e.g. primary care) or residual laboratory sera. Many studies undertaken quickly during the pandemic are under-powered to identify age-differences.

We undertook a systematic review and meta-analysis of published and unpublished literature to assess the susceptibility to SARS-CoV-2 in children and adolescents compared with adults. We limited this review to contact-tracing studies and population-based studies as these are likely to be most informative and least open to bias.

### Methods

Our review question was “What is the susceptibility to SARS-CoV-2 by children and adolescents compared with adults?”

We undertook a rapid systematic review and included contact tracing studies or prevalence studies in published or preprint form and including data from a national public health website reporting government statistics and studies. Studies were required to provide data on proven SARS-CoV-2 infection (PCR or serology) and report either rate of secondary infections in children and young people compared with adult or infection prevalence or seroprevalence in children and adolescents separately to adults.

We excluded reports of single household/institution outbreaks; studies of hospitalised patients, clinical studies and cohorts defined by symptoms; studies of unconfirmed cases i.e. cases based on self-report or symptoms, including contact-tracing studies where only symptomatic contacts were traced; modelling studies or reviews unless these reported new data; and prevalence studies with ascertainment based upon clinical contact and seroprevalence studies of residual sera, as these are likely to under-represent children

Where studies were drawn from populations that overlapped, we excluded studies where the time periods overlapped but included studies where time-periods did not overlap. We did not include in this review seroprevalence studies only in children as these did not allow comparison with adults.

We searched two electronic databases, PubMed and the medical preprint server medRxiv on 16 May 2020 and updated this on 28 July 2020. We used the following search terms in PubMed: (“COVID-19”[tw] OR “2019-nCoV”[tw] OR “SARS-CoV-2”[tw]) AND ((child* OR infant*) OR (“transmission”[tw] OR “transmission” [mh]) OR (“Disease Susceptibility”[tw] OR “susceptibility”(mh)) OR (“epidemiology”[tw] OR “epidemiology” [mh]) OR (“contact tracing”[tw] or “communicable disease contact tracing”[mh])). In medRxiv we undertook separate searches for ‘child and covid-19’, ‘covid-19 and epidemiology’, ‘covid-19 and susceptibility’ and ‘covid-19 transmission’ as more complex Boolean searches are not available.

Figure 1 shows the PRISMA flow diagram.

**Figure 1.**
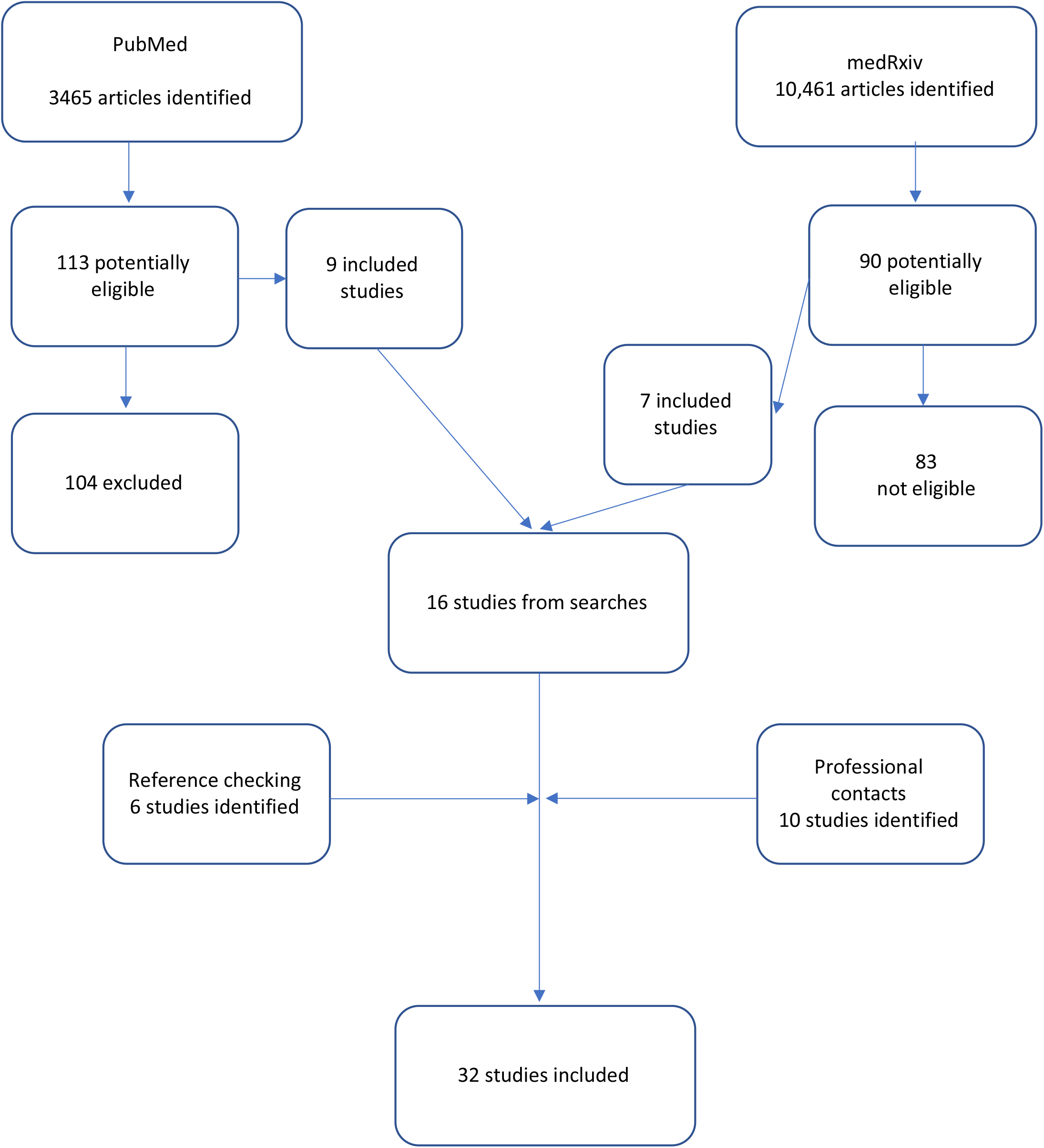
PRISMA flow diagram for search

One researcher (RV) screened studies on title and abstract to identify potentially eligible studies for full-text review. Full text studies were then reviewed by two researchers for eligibility and data were extracted independently by two researchers (RV and OM or CW). We hand-searched cited references in all potentially eligible studies for additional studies and identified additional studies through authors’ professional networks.

Data were extracted on country, study type, study context with regards social distancing measures and school closures at the time of the study, case definition, testing method, sampling method, and infection rates in adults and children.

Methodological quality of included studies was assessed independently by 3 authors (OM, CW, RV) based on a critical appraisal checklist for prevalence studies.^15^ We assessed risk of bias using two additional criteria: whether symptomatic contacts (in contact-tracing studies) or individuals (population-screening studies) were more likely to participate than asymptomatic ones; and whether the obtained sample was >75% of the intended sample. Studies were categorised as high quality if they met all quality criteria and had low risk of bias on both criteria; medium if they had low risk of bias on 1 or more criteria and met ≥5 of 7 quality criteria; low if they had met <5 quality criteria; or Uncertain if multiple domains could not be scored.

#### Analysis

Contact tracing and population prevalence studies were considered separately. Random effects meta-analysis with restricted maximum likelihood estimation was undertaken using the *meta* commands in Stata 16 (StataCorp; College Station, TX). Odds ratios were used as the primary metric for contact tracing studies. Prevalence ratios were used as the primary metric in population-based studies. We planned subgroup analyses using restricted maximum likelihood based upon quality of study and age of children / adolescents.

We followed the PRISMA guidelines in reporting findings.

No funding was obtained.

### Findings

The PubMed search resulted in 3465 and the medRxiv search in 10,461 studies, of which 113 and 90 respectively were examined in full text and 16 studies included (Figure 1). We identified a further 6 studies through reference-checking and 10 studies through professional networks. In total 32 studies were included (Table 1) with quality/bias assessments shown in Appendix Table 1. Eighteen were contact-tracing studies (CTS) (3 were school CTS), and 14 were population-screening studies. Two were high quality, 22 medium, 7 low and one uncertain.

**Table 1.** Characteristics of included studies

| A. Contact-tracing studies |  |  |  |  |  |  |  |  |
| --- | --- | --- | --- | --- | --- | --- | --- | --- |
| Author | Status | Location | Recruitment of index cases | Recruitment and isolation of contacts | Contact type | Number of clusters, index cases and contacts | Case definition/testing | Age: child / adult |
| Zhang et al.<sup>10</sup> | Published & peer reviewed | Hunan, China | All confirmed cases identified by Hunan CDC between 16 January and 1 March 2020. | January 16, 2020 to March 1, 2020. Close contacts were identified through contact tracing of a confirmed cases and placed under medical observation for 14 days. A close contact is defined as an individual who had unprotected close contact (within 1 meter) with a confirmed case or an asymptomatic infection within 2 days before their symptom onset or sample collection. | All contact types | 114 clusters representing 136 index cases & 7193 contacts.<br>]One (0.7%) index case was <15 years. | RT-PCR positive<br>All close contacts were tested in accordance with local policy regardless of symptoms. % of contacts tested not stated. | 0-14y / 15+y |
| Li et al.<sup>18</sup> | Published & peer reviewed | Hubei, China (Hospitals in Zaoyang City and Chibi City) | Index cases identified from two hospitals (in Zaoyang City and Chibi City) to 13 February 2020. Index cases were excluded if members of their family had links to Wuhan. Not clear if all cases from hospital were sampled or just a sub-set. | 1 January to 13 February 2020. All household contacts were quarantined immediately for 14 days by the local government and monitored daily. | Household contacts | 105 index patients with their households (n=105) and all family contacts (n=392).<br>The proportion of index cases who were children was not reported. | RT-PCR positive<br>Nasopharyngeal swab samples were collected at the beginning and the middle of quarantine. 100% of contacts tested 2-4 times. | 0-17y / 18+y |
| Cheng et al.<sup>20</sup> | Published & peer reviewed | Taiwan | The initial 100 confirmed cases in Taiwan between 15 January and 18 March 2020. | Close contacts were identified through epidemiological investigation and defined as a person who did not wear appropriate personal protection equipment (PPE) while having face-to-face contact with a confirmed case for more than 15 minutes during the investigation period (defined by epidemiological investigation and typically up to four days prior to symptom onset or test date for asymptomatic cases). All close contacts were quarantined at home for 14 days after their last exposure to the index case. | All contact types | 100 index cases; 2761 close contacts.<br>The youngest index case was age 11 years although the proportion of index cases that were children was not reported. | RT-PCR positive.<br>Routine testing for household and healthcare worker contacts (30.7%).<br>Other contacts (69.3%) were only tested if symptomatic. | 0-19y / 20+y |
| Wang et al.<sup>17</sup> | Published & peer reviewed | Wuhan, China | Patients hospitalized in Union Hospital (n=85) on 13 and 14 February. Not clear if all cases from hospital were sampled or just a sub-set. | Household contacts of the hospitalised patients, followed for 14 days. | Household contacts | They enrolled 85 households corresponding to the 85 patients and identified 155 household contacts. | RT-PCR positive<br>Throat swabs. Process for testing household members not stated, but 33% of household contacts were not tested for SARS-CoV-2 | Child age not defined. |
| Mizumoto et al.<sup>53</sup> | Preprint | Japan | Cases that were domestically acquired and confirmed by RT-PCR by 7 March 2020 | Contacts of index cases, definition and method of ascertainment not given. No details on isolation of contacts. | Not stated. the total number of contacts (8 per index case) suggests these are likely all contacts. | 313 cases and their 2496 close contacts. | RT PCR positive. Process and eligibility for testing of contacts not described. | 0-19y / 20+y |
| Wang, Tian et al.<sup>19</sup> | Published & peer reviewed | Beijing, China | All laboratory-confirmed (RT-PCR) cases in Beijing up to 21 February 2020, recruited through Beijing CDC. | 28 February and 8 March 2020. All household members of index cases were followed for 14 days. Testing and quarantine of contacts not clearly defined. | Household contacts | 124 of 137 eligible families participated.<br>No primary cases were <18y. | Index and secondary cases defined by RT-PCR positive. Proportion of PCR testing of secondary contacts is not stated. |  |
| Park et al.<sup>12</sup> | Published and peer reviewed | South Korea | All laboratory-confirmed cases in Korea registered with Korea CDC from 20 January to May 13. | All contacts of index cases registered with Korea CDC through a comprehensive national contact-tracing system and followed for a mean of 10 days. | Household and non-household contacts. Note only data on household contacts included in this review. | Studied 59,073 contacts (10,592 were household contacts) of 5706 index cases. Only included index cases who reported 1 or more contact however only included 52% of 10,962 national cases reported in the period. | Household and healthcare worker contacts routinely tested by RT-PCR. Other contacts only tested if symptomatic. |  |
| Dattner et al.<sup>24</sup> | Preprint | Israel | Identification of all households in city of Bnei Brak where all household members had been tested (PCR) and 1 or more member was positive. Households identified through the Israeli COVID-19 database until 2 May 2020. | All household members included.<br>Note 51% of population is < age 20 years. | Household | 637 houses comprising 3353 people of whom 1510 were positive. All eligible households were included.<br>The figures included in our systematic review were derived from supplied estimated probabilities of children or adults being the index. | RT-PCR testing of all household members including index cases and contacts. |  |
| Hu et al.<sup>16</sup> | Preprint | Hunan, China | All cases with contact details were identified from the notifiable infectious diseases reporting system in Hunan Province. 16 Jan to 2 April 2020. | Contacts were quarantined for 14 days and tested (PCR) at least once during quarantine: after 7 Feb all contacts were tested but only symptomatic contacts tested before 7 Feb (approx. 50% of contacts tested). | All contacts | 1178 cases and their 15,648 contacts. | RT-PCR |  |
| Laxminarayan et al.<sup>28</sup> | Preprint | Tamil Nadu & Andhra Pradesh, India | Index cases identified from state registries and contacts traced by public health agencies in each state- 5 March to 4 June (to 29 May in A.P). | Contacts traced by public health agencies and tested between 5-15 days of exposure. Insufficient detail provided. Note that there were twice as many close contacts per index case <18y compared with >18y. | All contacts | 4206 confirmed cases and 64,031 contacts.<br>Note only 4206 cases included out of 33,584 total cases =13%, with no detail given on non-recruitment. | RT-PCR of all contacts regardless of symptoms. |  |
| Liu et al.<sup>26</sup> | Published and peer reviewed | Guandong Province, China | All cases identified by intensive regional surveillance by local CDC from 15 Jan to 15 March | Contacts traced and monitored with PCR from throat swabs taken every few days for 14 days; 84% of contacts were quarantined in centralised stations. | All contacts | 1361 cases reported and 11,868 contacts traced and quarantined. Analysis included 11,580 contacts (98%). | RT-PCR from throat swabs |  |
| Rosenberg et al.<sup>22</sup> | Published and peer reviewed | New York State (excluding New York City), USA | Identified and studied 229 initial confirmed (PCR) cases in NY State outside of NY City from 2 to 12 March. | Active contact tracing by county and state health departments. All household contacts were eligible for PCR testing. Contacts tested 0-10 days after index case (43% were tested on Day 0 i.e. initial index diagnosis day). | Household | 229 index cases and 343 household contacts. | RT-PCR<br>All household contacts were eligible for PCR testing, however not stated what proportion were tested. |  |
| Yousaf et al.<sup>23</sup> | Published and peer reviewed | Milwaukee (Wisc) and Salt Lake City (Utah), USA | All PCR-positive cases from two cities were identified through routine public health surveillance and recruited between 22 March and 22 April. | Active contact tracing by public health departments. All contacts were observed for 14 days with two swab tests (RT-PCR) on day 0 and day 14 plus if symptomatic. | Household | 195 of 198 contacts participated (98.5%).<br>Numbers of index cases not stated. | RT-PCR<br>All household contacts tested. |  |
| Chaw et al.<sup>27</sup> | Preprint | Brunei | All 71 initial cases in Brunei, which arose following a religious event, with cases detected after 9 March 2020. | Detailed contact tracing by Ministry of Health, with RT-PCR testing of all reported contacts. All contacts were quarantined for 14 days and retested if symptomatic. | All contacts | 71 index cases and 1755 close contacts. All contacts participated. | RT-PCR |  |
| Van der Hoek et al.<sup>7,25</sup> | Published and peer reviewed | Netherlands | National surveillance data from two Dutch systems<br>A. Osiris: registry of all laboratory-confirmed cases<br>B. HP Zone: data on contact tracing from 23 of 25 Dutch municipalities (GCDs). | Data included to 2 April.<br>Contact-tracing was undertaken for all cases registered in HPZone. Contact infection status identified through linkage to the main national surveillance database, suggesting that only symptomatic secondary cases were included. | All contacts | 231 cases and 709 close contacts.<br>Proportion of contacts tested not stated | RT-PCR | <19y |
| **B. School contact-tracing studies** |  |  |  |  |  |  |  |  |
| Macartney et al.<sup>29</sup> | Published and peer reviewed | New South Wales, Australia | COVID-19 cases in 25 educational settings (15 schools & 10 early learning centres) for which a person (student or staff) with proven COVID-19 (PCR positive) had attended while infectious. Identified through state Notifiable Conditions Information Management System. Schools remained open but | 25 Jan to 9 April 2020. Followed up all close contacts (a person who has been in face to face contact for at least 15 minutes or in the same room for at least 40 minutes with a case while infectious). All close contacts followed and tested if symptomatic during the 14 day isolation period. | Educational setting contacts only | 27 primary cases (12 student; 15 staff cases) and their 1448 school/early learning-related close contacts from 25 educational settings.<br>12 high school cases (8 students; 4 staff) from 10 | RT PCR or serology (specific IgG, IgA, IgM detection using indirect immunofluorescence) positive.<br>Swabs taken from | 6w-18y / 20y+ |
|  |  |  | students dismissed from 23 March (<5% student attendance). Note that school attendance remained high at the time that secondary cases were identified in schools, and early-years settings did not close. | 7 settings had testing of all contacts 5-10 days after last contact plus serology after day 21. |  | schools had a total of 695 contacts (598 students; 97 staff). The 5 primary school cases (1 student; 4 staff) from 5 schools had a total of 218 contacts (179 student; 39 staff) 1448 contacts identified; 663(43.5%) were tested (PCR or serology or both). | 542/1,448 contacts (37.4%). Serology was performed in 208/1448 contacts (14.3%). |  |
| Heavey et al.<sup>30</sup> | Published and peer reviewed | Republic of Ireland | Screened the Republic of Ireland national surveillance to identify all PCR-positive cases in children or adults who attended school settings in period before schools were closed on March 12 2020. | 1-12 March 2020. Contacts traced and advised to quarantine at home for 14 days. Tested (PCR) only if symptomatic. | All contacts including school contacts | 6 index cases identified (3 adult; 3 <18y).<br>1155 contacts identified (924 child; 101 adult). | RT-PCR testing if symptomatic | 0-17y / 18y+ |
| Yung et al. | Published and peer reviewed | Singapore | 3 potential SARS-CoV-2 seeding incidents in educational settings in Singapore identified from national surveillance during February and March 2020. | Feb to March 2020. Close school contacts (e.g. classmates) quarantined for 14 days. Contacts in 1 school and 1 preschool were tested only if symptomatic; these schools were not closed. Contacts in 1 preschool were tested (PCR) after an outbreak causing school closure. | School contacts only | Three PCR-positive child index-cases were identified from 2 preschools and 1 secondary school.<br>188 contacts studied, of whom 119 (63%) were tested. | RT-PCR | 1-16y |

| C. Population-screening studies |  |  |  |  |  |  |  |  |
| --- | --- | --- | --- | --- | --- | --- | --- | --- |
| Author | Status | Location | Context | Recruitment | Timing of survey | Note | Case definition/testing | Age: child / adult |
| Gudbjartsson et al.<sup>32</sup> | Published & peer reviewed | Iceland | First infection diagnosed on 28 February 2020; Containment measures put in place. Primary schools open but some secondary schools closed and moderate restrictions on social contacts from 13 March. | 13 March to 6 April 2020. National population screening. Open invitation for 87% of participants through online portal but with collection of sample from one location (Reykjavik), and random invitation for a sub-sample (13%). Children <10y made up 6.4% of sample.<br>Participation in the study was primarily by request of participants rather than by random sampling, which may have introduced biases in participation. | 13 March to 6 April. | Only population-screening sample reported here. | RT-PCR on nasopharyngeal and oropharyngeal samples. | 0-9y / 10+y |
| Lavezzo et al.<sup>34</sup> | Published & peer reviewed | Vo, Veneto Region, Italy | Quarantined community in an area of Italy that was affected early and severely | All age groups were homogeneously sampled with age-specific percentages ranging from 70.8% to 91.6%. Two surveys undertaken; first survey only | 21-29 February 2020 | We present data only from this first survey although the paper also reports a | RT-PCR on nasopharyngeal samples. | 0-20y / 21+y |
| Author | Status | Location | Context | Recruitment | Timing of survey | Note | Case definition/testing | Age: child / adult |
|  |  |  | in the epidemic; area was 'locked down' from the 23 February for two weeks. Study undertaken close to the imposition of very strict social distancing measures in the region. | included here (overall response rate 85.9%). Those <21y made up 17% of sample and had a participation rate of 94% (0-10y) and 95% (11-20y) |  | second survey undertaken during 'lockdown'. |  |  |
| Swedish National Study<sup>33</sup> | Online report | Sweden | First death reported in Stockholm on 11 March 2020. Voluntary social distancing measures recommended from 16 March 2020, with secondary schools recommended to teach virtually. Primary schools and early years settings remained open throughout. | Two nationally-representative surveys undertaken by the Swedish Public Health Agency, Folkhälsomyndigheten.<br>Participants invited by email: 25 71/4480 (57%) participated in April and 295 7/4487 (66%) in May.<br>Children 0-15y made up 18.9% of the April and 17.2% of the May sample<br>Participants performed home self-sampling using nasopharyngeal swabs. | 21-24 April and 25-28 May, |  | RT-PCR on nasopharyngeal samples. | 0-15y / 16+y |
| UK ONS<sup>35</sup> | Online report | England | Strict national social distancing measures enacted 20 March 2020, with gradual easing of lockdown from 25 May. | Representative sample of 35,801 individuals in England. Those 2-19y made up 17% of the population. Cases were identified by home self-sampling using nasopharyngeal swabs with carers swabbing young children.<br>79% of invited participants provided 1 or more swabs. | 26 April-27 June 2020 | Repeated surveys carried out each week. Data shown here are the cumulative prevalence of those ever positive between 26 April-27 June. | RT-PCR on nasopharyngeal samples. | 2-19y / 20+y |
| Pollan et al. ENE-COVID-19<sup>36</sup> | Online report | Spain | Strict social distancing was imposed on 14 March 2020. Some restrictions were lifted on 27 April and further restrictions lifted on 11 May. | Undertaken by Spanish Ministry of Health. National representative sample obtained from random sampling of households in municipalities across Spain. 61,075 participants provided point of care samples (59%) and 51,958 included in both immunoassay and point of care - out of 102,803 approached<br>Those 0-19 years (n=11,464) made up 23% of the point of care sample and 12.6% of the immunoassay sample. | 27 April - 11 May 2020 | We used the point of care data here due to the sample being representative of the child population, unlike the immunoassay test. | Point of care test: rapid immunochromatography IgG: Orient Gene, Zhejiang Orient Gene Biotech.<br>Immunoassay: Abbott IgG serology.<br>Comparison of the rapid test IgG with SARS-CoV-2 serology in 16,953 of the study sample found 97.3% agreement between tests. | 0-19y / 20+y |
| Author | Status | Location | Context | Recruitment | Timing of survey | Note | Case definition/testing | Age: child / adult |
| Netherlands Pienter<sup>7</sup> | Online report | Netherlands | Social distancing measures introduced gradually from 11 March 2020. Schools closed from 15 March. | Undertaken by the Netherlands National Institute for Public Health and the Environment (RIVM). Population-based sampling was undertaken in a random sample of a randomly chosen subset of municipalities across the Netherlands. Total sample of 2096. Those <20y made up 20% of sample. | 31 March - 13 April 2020 | Data provided by author FdK. | Serology (IgG) | 0-19y / 20+y |
| Hallal et al.<sup>42</sup> | Preprint | Brazil | First cases reported 27 February with local/state lockdowns during March and April. Some states began to relax measures in April. | Nationwide seroprevalence survey in 133 sentinel cities in 26 Brazilian states. Randomly selected households visited and finger-prick rapid serology test used. Total sample was 24,995 with household response rate =55%. Children heavily under-represented – 0-9y were 2.2% and 10-19y were 9.1%. | 14-21 May 2020 |  | Rapid lateral flow test used in our analysis (Wondfo SARS-CoV-2). | 0-19y |
| Shakiba et al.<sup>38</sup> | Preprint | Iran | Population-based seroprevalence study in 5 counties in Guilan province, northern Iran in April 2020 – previously very high virus prevalence. | multi-stage cluster random sampling approach and telephone recruitment of head of household. 196 /632 approached households participated (31%) – with n=528 participants. | April until 23 April 2020 |  | VivaDiag COVID 19 IgM/IgG serology. |  |
| Biggs et al.<sup>39</sup> | Published and peer reviewed | Georgia, USA | Study undertaken by US Centers for Disease Control (CDC) to coincide with end of shelter in place orders (3-30 April). | Survey of a random sample of households in two metropolitan Atlanta counties. 696 persons from 394/1675 households (23.5%) participated. Children <18y were 6.9% of sample compared with 22.4% of population. | 28 April – 3 May 2020 |  | Total antibody measured using VITROS 3600 Immunodiagnostic System (Ortho Clinical Diagnostics). | 0-17y / 18y+ |
| Stringhini et al.<sup>40</sup> | Published and peer reviewed | Geneva canton, Switzerland | First case on 26 Feb 2020. Schools closed on 16 March and strict social distancing measures introduced 20 March. Seroprevalence initiated using a population-based sample in canton. | Population-based but not fully random sample within canton (region). 1300 randomly selected adults approached each week for 5 weeks and invited to bring all household aged 5+ for serology. Only non-symptomatic individuals studied. 2766/5492 (50.4%) agreed to participate in total, and data presented here for first 1360. 16.4% of sample aged 0-19y, similar to population. | 6 April – 9 May 2020 | Indeterminate cases were treated as negative in calculating data for the meta-analysis. | ELISA to spike protein (Euroimmun; Lübeck, Germany #EI 2606-9601 G) | 5-19y / 20+y |
| Nawa et al.<sup>41</sup> | Preprint | Utsunomiya City, Greater Tokyo, Japan | First cases in Japan from 15 January. All schools closed 27 February. Survey conducted | Population-based seroprevalence survey: a random sample of 1000 households approached 742/2290 persons (32%) participated. 13% were <18y – similar to population. | 14 June-5 July |  | IgG (Shenzhen YHLO Biotech Co., Ltd., Shenzhen, China). | 0-17y, 18y+ |
| Author | Status | Location | Context | Recruitment | Timing of survey | Note | Case definition/testing | Age: child / adult |
|  |  |  | between the first and second spikes of infection in the city. |  |  |  |  |  |
| Pagani et al.<sup>43</sup> | Preprint | Lombardy, Italy | The town of Castiglione d'Adda, 4550 inhabitants had high numbers of infections from early in the pandemic. Local lockdown occurred from 23 February 2020. | Entire population (all ages) invited to participate: recruited 4174 /4550 inhabitants (92%) who had rapid capillary testing, of whom a random sample of 562 (stratified for age and sex) had formal serology by venepuncture.<br>0-19y made up 12% of the rapid and formal serology samples. | June 2020 | 22% of population showed overall positivity (22.2% on rapid test, 22.6% on formal serology)<br>Rapid test used in meta-analyses here as findings from formal serology were highly similar. | Rapid capillary testing: lateral-flow immunocromatographic test (Prima Lab, Switzerland)<br>Serology: CLIA, IgG anti-SARS-CoV-2, Abbott, USA), | 0-19 / 20+ |
| Weis et al.<sup>44</sup> | Preprint | Thuringia, Germany | Seroprevalence survey (CoNAN study) in in the previously quarantined community Neustadt-am-Rennsteig, from six weeks after a SARS-CoV-2 outbreak (March 22<sup>nd</sup>). Local lockdown initiated. | All community households invited. Enrolled 626/883 = 71% of community.<br>Focus on child participation and blood collection to be representative. Children 1-17y were 9.5% of the sample<br>620 gave blood and 600 participants had all 6 serological tests performed. | 12-16 May 2020 |  | Serology by 6 quantification methods: 2 ELISA and 4 immunoassay. EDI Novel Coronavirus SARS-CoV-2 IgG; ELISA kit (Epitope Diagnostics Inc., San Diego, USA), SARS-CoV-2 IgG ELISA kit ; (Euroimmun, Lübeck, Germany), SARS-CoV-2 S1/S2 IgG CLIA kit (DiaSorin, Saluggia, Italy), ; 2019-nCoV IgG kit (Snibe Co., Ltd., Shenzhen, China), SARS-CoV-2 IgG CMIA kit (Abbott); Chicago, USA) and Elecsys Anti-SARS-CoV-2 kit (Roche, Basel Switzerland). | 1-17 / 18+ |
| Streeck et al.<sup>45</sup> | Preprint | Gangelt, Germany | Carnival held on 15 February. Strict local social distancing measures introduced on 28 February due to local outbreak and deaths. | A random sample of 600 households was invited to participate and 1007 individuals from 405 households participated. 919 provided serology data. 5-14y olds made up 6.0% of sample. | 30 March – 7 April 2020 | 62% of the 88 participants who could not be assessed were children not assessed for technical reasons. | Serology (IgG) | 5-14y / 15+y |

#### Contact tracing studies

Six were from mainland China, two from the USA and one each from Taiwan, Japan, South Korea, Israel, the Netherlands, Brunei and India, with school CTS from Australia, the Ireland and Singapore. Lower secondary attack rates (SAR) in children and young people compared with adults were reported by 11 studies; 5 from provinces of China, including Hunan^10,16^ Hubei,^17,18^ and Beijing;^19^ and 6 studies from other countries, including Taiwan,^20^ Japan,^21^ the USA,^22,23^ Israel^24^ and the Netherlands,^7,25^ although confidence intervals were wide in some studies.

No significant differences in SAR by age were reported in four studies: from Guangdong province, China,^26^ Brunei^27^ and the states of Tamil Nadu and Andhra Pradesh in India^28^ with one study from South Korea reporting high SAR in <19 year-olds.^12^ In three of these, SAR in younger children were low compared with adults but those amongst teenagers were as high as or higher than adults. ^12,27,28^

We undertook a random effects meta-analysis of SAR in children and young people compared with adult, with data able to be included from 14 studies. We combined data on children and young people <20 years and adult age-groups >20 years, thus odds ratios (OR) and prevalence rates for adults may differ from those reported in studies. The pooled OR estimate for all contact-tracing studies of being a child with secondary infection compared with adults was 0.56 (0.37, 0.85) with high heterogeneity (95%) (Figure 2).

**Figure 2.**
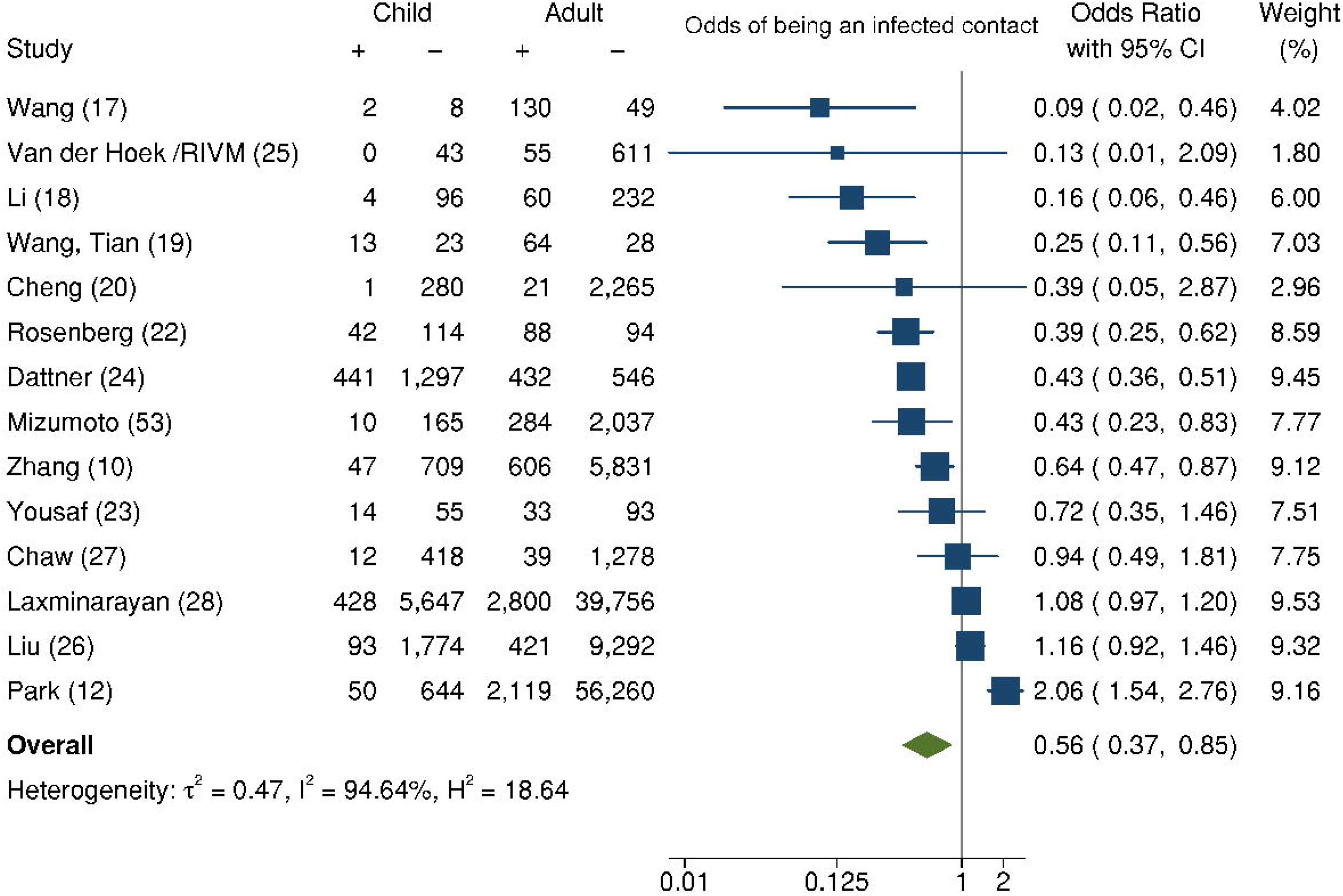
Pooled estimate of odds of being an infected contact among children compared with adults for all contact tracing studies

In meta-analysis of 8 CTS grouped by age of child (Figure 3), the pooled OR for children <12-14 years was 0.52 (0.33, 0.82), significantly lower than adults, whereas for adolescents this was non-significant (OR=1.23 (0.64, 2.36). Chi-square test suggested this group difference was significant (chi-2 =4.54, p=0.033).

**Figure 3.**
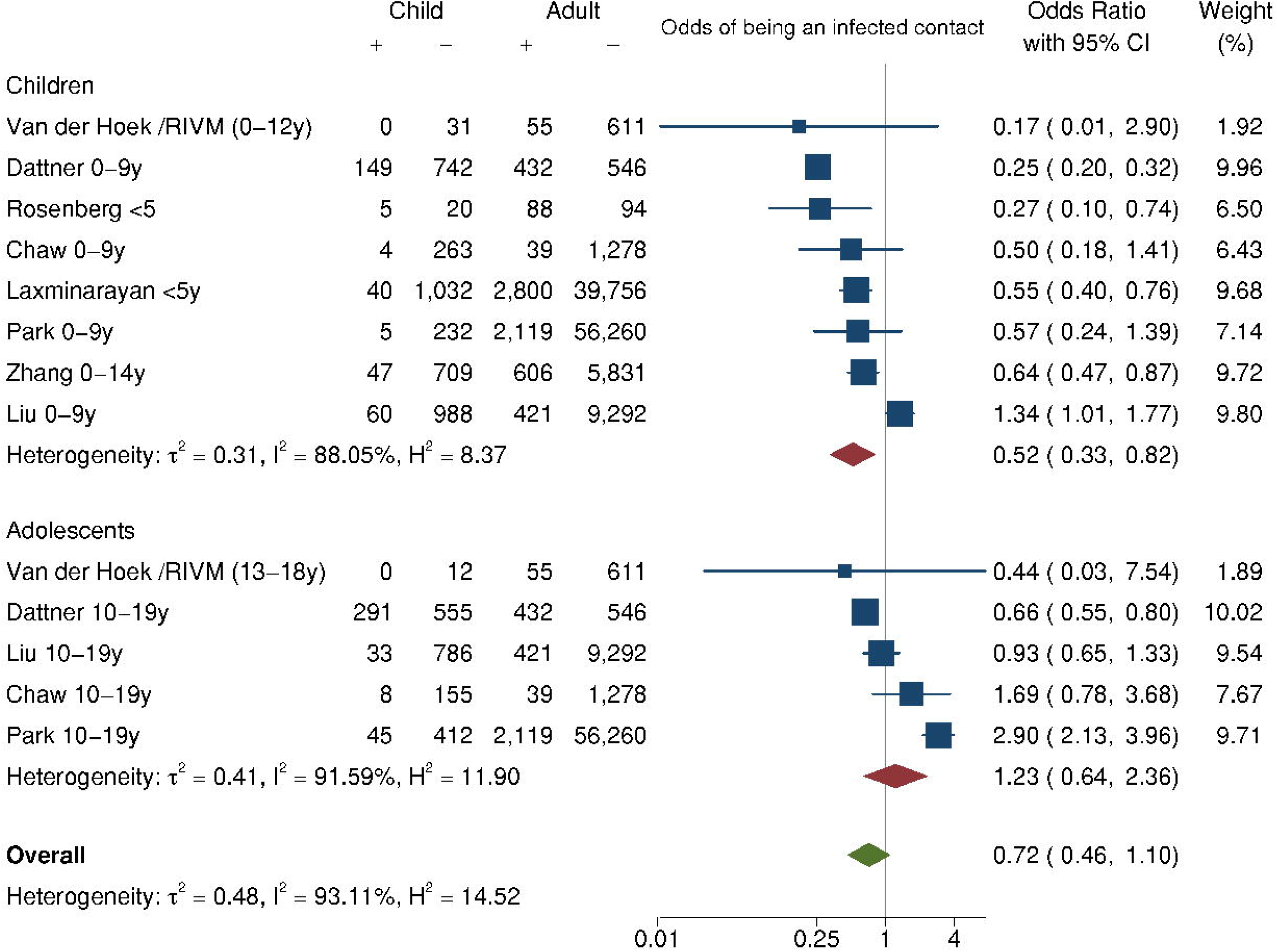
Pooled estimate of odds of being an infected contact among children and among adolescents compared with adults for contact tracing studies

When only the 8 medium/high-quality (low risk of bias) studies were examined, this finding was no longer significant (OR 0.68 (0.41, 1.11), however the difference in estimates between low and medium/high quality studies was not significant (p=0.202). (see Appendix Figure 1).

We hypothesised that CTS including only household contacts might provide a clearer indication of the relative susceptibility to infection of children versus adults because all contacts within households might be assumed to receive a similar exposure to infection from index cases. A post-hoc analysis by type of contacts (Appendix Figure 2) showed studies of household contacts gave a lower pooled odds ratio (0.41 (0.22, 0.76)) than did studies of all contacts (0.91 (0.69, 1.21)) (between group variance; df=l, chi2= 5.31, p=0.021).

Three studies undertook contact-tracing in schools. A state-wide population-based CTS in educational settings in Australia before and during school closures^29^ found that 27 primary cases (56% staff) across 25 schools or early-years nurseries resulted in 18 secondary cases in 4 settings, including an outbreak of 13 in one early-years setting initiated by a staff member with no evidence of child to adult transmission. The SAR was 1.2% (18/1448) overall, 5/1411=0.4% excluding the early-years outbreak and 2.8% (18/633) in those tested. Other national CTS undertaken in schools in the Republic of Ireland^30^ and Singapore^31^ before schools closed identified very few secondary cases in schools.

#### Population screening studies

Data from prevalence studies for children and young people compared with adults is shown in Figure 4. We did not undertake a meta-analysis of population-screening studies, given the important differences in the populations, epidemic time-points and methodologies involved.

**Figure 4.**
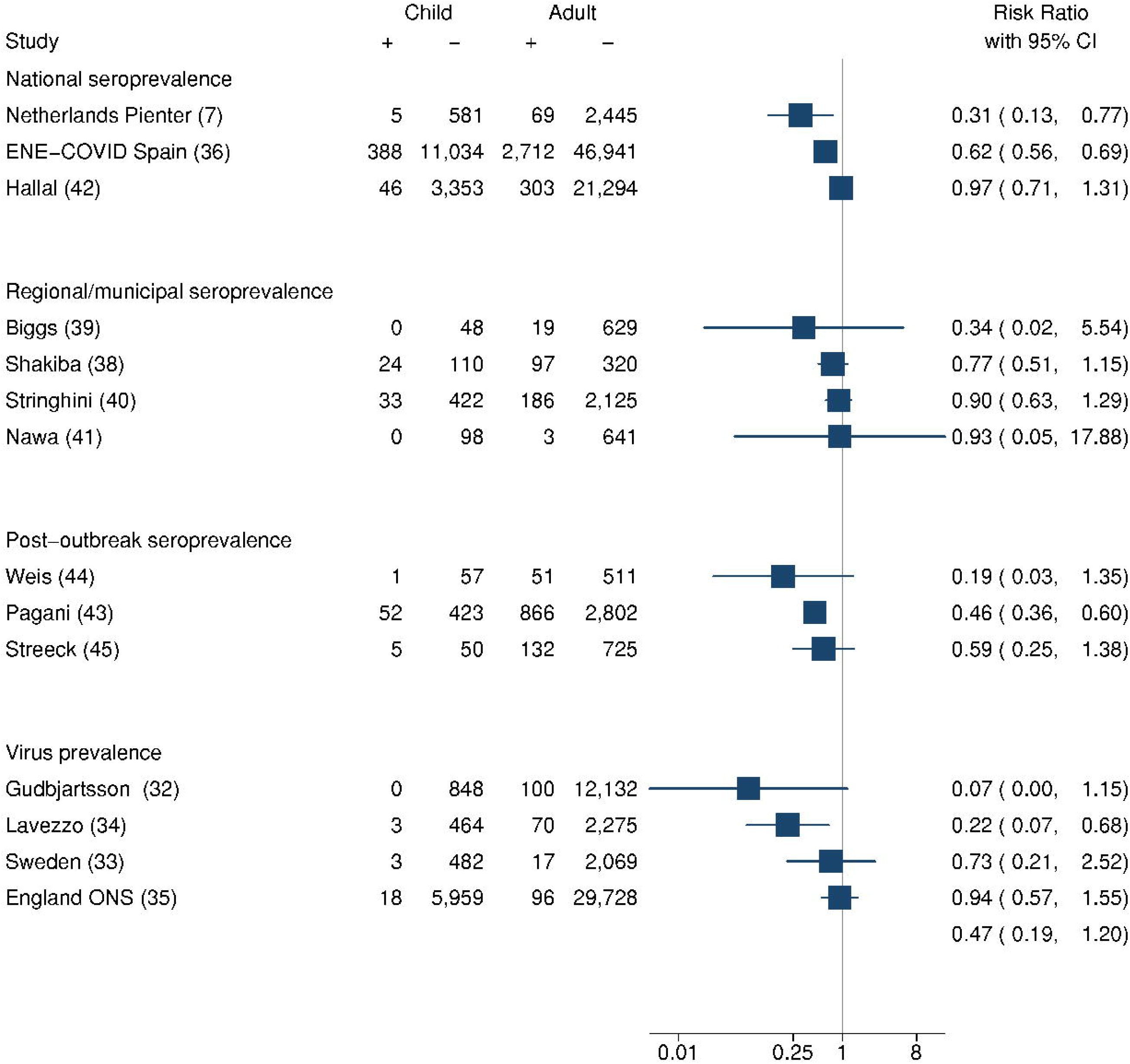
Ratios of the prevalence of SARS-CoV-2 infection in children and young people compared with adults in population-screening studies

Four studies reported virus prevalence. National prevalence studies from Iceland^32^ and Sweden^33^ undertaken while primary schools were open, showed lower prevalence amongst children and young people than adults, as did a municipal study from Italy^34^ undertaken just before lockdown while schools were open. However a nationally-representative survey from England covering lockdown and the subsequent month identified no significant differences by age.^35^

10 studies reported seroprevalence, 3 being nationally representative. A lower seroprevalence was identified in children and in some instances adolescents compared with adults in a number of studies, including a nationally representative study in Spain (ENE-COVID-19),^36^ a Dutch nationally-representative study (Pienter Corona study),^7,37^ and city or regional studies from Iran,^38^ the USA,^39^ Switzerland^40^ and Japan^41^ although no difference by age was found in a survey in 133 sentinel cities in 26 Brazilian states.^42^ Two community-based studies following localised outbreaks found lower seroprevalence amongst children and young people than adults in Lombardy, Italy^43^ and Thuringia, Germany,^44^ with a second German post-outbreak study finding no overall association with age.^45^

Examination of seroprevalence findings in children separately to adolescents (Appendix Figure 3) suggested that seroprevalence lower than adults amongst younger children (<10 years) but not in adolescents, although this was not formally tested.

## Discussion

We identified 37 studies from 23 countries that met our eligibility criteria and provided information on susceptibility to and transmission of SARS-CoV-2 in children and young people compared with adults. We excluded studies and study types open to very significant bias, yet studies were predominantly of medium and low quality, with only two high quality studies. The majority of studies were from middle and high-income countries in East Asia and Europe.

We found preliminary evidence from 15 contact-tracing studies that children and young people have lower susceptibility for SARS-CoV-2 infection than adults, with a pooled odds ratio of 0.57 (0.39, 0.83). This estimate was little changed when only medium or high quality studies were examined, although power was reduced and the confidence interval included one. Only one study^13^ found a higher odds of infection in 0-19 year olds than adults, although this finding was confined to 10-19 year olds. When studies were categorised by age of the children, lower susceptibility appeared to be confined to younger children (less than 14 years), who had a 48% lower odds of infection compared with adults aged ≥20 years. The age bands of the studies were not aligned making direct comparisons challenging.

Data from population-screening studies were heterogenous and were not suitable for meta-analysis. Findings consistent with lower seroprevalence in 0-19 year olds compared with adults were reported by two national studies, one regional study and all of the municipal post-outbreak studies, although confidence intervals were wide in some cases. Two virus prevalence studies similarly reported lower infection rates in ≤20 year-olds. In contrast, other studies reported no age-related differences. No studies reported higher prevalence in children and adolescents. Examination of seroprevalence findings in children separately to adolescents showed that the majority of studies were consistent with lower seroprevalence in children compared with adults, although seroprevalence in adolescents appeared similar to adults in all studies.

The findings from the CTS and prevalence studies are largely consistent in suggesting that children below approximately 12-14 years are less susceptible to SARS-CoV-2 infection, resulting in lower prevalence and seroprevalence than adults. Data specifically on adolescents are sparse although consistent with susceptibility and prevalence more similar to adults. Our findings on susceptibility are similar to a modelling analysis by Davies et al.,^46^ who estimated that those under 20 years were approximately half as susceptible to SARS-CoV-2 as adults.

We found few data that were informative on the onward transmission of SARS-CoV-2 from children to others. Data from the large Australian school contact-tracing study suggest that, at a population level, children and young people might play only a limited role in transmission of this pandemic. This is consistent with the data on susceptibility noted above, i.e. suggesting that lower rates of secondary infection mean that children and young people have less opportunity for onward transmission. There is evidence of transmission from children to others in households and in schools, and there have been reported outbreaks in schools.^47,48^ Other very small studies in Ireland^30^ and Singapore^31^ have found low numbers of secondary cases resulting from infected children attending school. This is consistent with a national South Korean study, which found the SAR from children to household members was extremely low.^13^ The available studies suggest children and young people play a lesser role in transmission of SARS-CoV-2, in marked contrast to pandemic influenza.^49^

### Limitations

Our study is subject to a number of limitations. We remain early in the pandemic and data continue to evolve. It is possible that unknown factors related to age, e.g. transience of infection or waning of immunity, bias findings in ways we don’t yet understand. Some studies were low quality and nearly all included studies were open to bias. The secondary infection rate in some CTS was low and this may represent an underestimate of the unmitigated household attack rate of SARS-CoV-2 as transmission chains were cut short because of strict control measures.^50^ Most of the CTS were undertaken when strict social distancing measures had been introduced, e.g. closures of schools and workplaces, restriction of travel. This would have reduced contacts outside the home, especially contacts between children, but it may have increased contacts between children and adults by increasing the household contact rate. The number of contacts nominated and traced for 0-19 year olds was low compared with adults in some studies,^12,28^ which may have introduced bias. We identified 3 CTS from Guangdong province^11,51,52^ which were excluded as they overlapped with Liu et al.,^26^ however findings were unchanged if these studies were included. We included two recent large CTS from India^28^ and South Korea^12^ however numbers of children and data quality appeared low, making firm conclusions difficult.

For population screening studies, the numbers of children tested was small in most of the studies, and was frequently less than the 15-25% of the population that are < 18 years in most countries. This likely reflects lower recruitment of children and may be a source of bias, although the direction of this bias is unclear. Age-differentials in sensitivity of swab or antibody tests may also confound findings. Interpreting the observed prevalence and seroprevalence studies requires thorough quantification of social mixing and transmission between age groups and how that changed during lockdowns and social distancing interventions.

### Summary and implications

There is preliminary evidence that children under 12-14 years have lower susceptibility to SARS-CoV-2 infection than adults, with adolescents appearing to have similar susceptibility to adults. There is some weak evidence that children and young people play a limited role in transmission of SARS-CoV-2 however this is not directly addressed by our study.

We remain early in our knowledge of SARS-CoV-2 and further data are urgently needed, particularly from low-income settings. These include further large, high quality contact-tracing studies with repeated swabbing and high-quality virus-detection and seroprevalence studies. Studies which investigate secondary infections from child or adolescent index cases in comparison to secondary infections from adult index cases are particularly needed in order to assess transmission. Monitoring of infection rates and contact-tracing studies within child-care and school settings will also be important. A range of serological studies are planned in many countries and these need to be sufficiently powered to assess differences in seroprevalence across different age groups and include repeated sampling at different time periods as social distancing restrictions are lifted. We will continue to update this review, including further data as available and updating preliminary data from some included studies.

## Data Availability

All data available from included studies, all available as open access

## Declarations

### Ethics

No ethical approvals were required for these secondary analyses of existing datasets.

### Consent for publication

Not applicable

### Access to data

Russell Viner had full access to all the data in the study and takes responsibility for the integrity of the data and the accuracy of the data analysis.

### Availability of data and materials

All included articles are on public access – see Appendix Table 1 for hyperlinks.

### Author’s contributions

RV and RME conceptualised the review. RV developed the search terms with the assistance of JT. RV undertook the initial searches. Data extraction was undertaken by RV, RE and OM. Quality assessment was undertaken by OM, CW and RV. Meta-analyses were done by RV with input from GM-T and JT. FvdK supplied additional data from one study. RV, CB and RME led the writing of the paper. All authors contributed to editing the paper and approved the final manuscript.

